# Immunosequencing of the T-Cell Receptor Repertoire Reveals Signatures Specific for Identification and Characterization of Early Lyme Disease

**DOI:** 10.1101/2021.07.30.21261353

**Authors:** Julia Greissl, Mitch Pesesky, Sudeb C. Dalai, Alison W. Rebman, Mark J. Soloski, Elizabeth J. Horn, Jennifer N. Dines, Darcy B. Gill, Rachel M. Gittelman, Thomas M. Snyder, Ryan O. Emerson, Edward Meeds, Thomas Manley, Ian M. Kaplan, Lance Baldo, Jonathan M. Carlson, Harlan S. Robins, John N. Aucott

## Abstract

Highly specific T-cell responses play key roles in pathogen clearance and maintaining immunologic memory. Next-generation sequencing of the T-cell receptor (TCR) repertoire is an emerging diagnostic technology that capitalizes on the specificity of T-cell responses to probe pathogen exposure. The spirochete Borrelia burgdorferi (Bb) wields an array of antigens with dynamic and complex immunogenic potential, and application of TCR immunosequencing to characterize Bb infection presents opportunities to improve detection of early Lyme disease (LD). By immunosequencing TCR repertoires in blood samples from 3 independent cohorts of patients with early LD and controls from Lyme-endemic/non-endemic regions, we identified 251 public, LD-associated TCRs. These TCRs were used to train a classifier for detection of early LD. The classifier identified LD with 99% specificity and showed 1.9-fold higher sensitivity (56% vs 30%) compared with standard two-tiered testing (STTT). TCR positivity predicted subsequent seroconversion in 37% of STTT-negative patients, suggesting that the T-cell response is detectable before the humoral response. Higher TCR scores were associated with clinical measures of disease severity, including abnormal liver function tests, disseminated rash, and number of symptoms. A subset of LD-associated TCRs mapped to Bb antigens, supporting specificity of this approach. These results suggest that TCR testing may be a highly specific and sensitive approach for identifying LD, particularly in the initial days of illness.

## INTRODUCTION

Lyme disease (LD) is the most common tick-borne illness in the United States, with more than 450,000 estimated new cases annually (1–3). In the United States, LD is initiated by infection with the spirochetal bacterium *Borrelia burgdorferi (Bb)* transmitted from infected *Ixodes* ticks (1). In the days to weeks after the initial tick bite, symptoms may include a characteristic erythema migrans (EM) rash and nonspecific flu-like symptoms while signs and symptoms of disseminated infection can affect the joints, nervous system, heart, or other areas of the skin (4).

Presence of EM rash in Lyme-endemic areas is highly suspicious for LD and warrants immediate treatment without further testing. However, guidelines recommend serologic testing to support a diagnosis of LD for individuals with absent or atypical EM or suspected disseminated infection (5, 6). The most common laboratory test for LD is standard two-tiered testing (STTT), which combines an initial enzyme-linked immunosorbent assay (ELISA) with a more specific immunoblot for positive or equivocal samples to detect antibodies against *Bb* (7, 8). However, STTT has some notable limitations, including poor sensitivity (25-50%) in the very early stages of infection, when most serologic testing is conducted (9, 10). This is likely attributable to the kinetics of the humoral response, as sensitivity of STTT improves with time, potentially exceeding 99% in untreated individuals with later-stage disease (11). Up to 60% of individuals testing negative within the first days to weeks after onset of symptoms may test positive upon re- testing 30 days later (10, 12–17). Additionally, false positives and interlaboratory variability may occur with STTT due to poor specificity and weak immunoreactivity (18, 19). These limitations underscore the unmet clinical need for improved methods for diagnosing LD, especially in the early stages of infection.

Infection with *Bb* elicits a T-cell response with kinetics that differ from the humoral response (20–22). Evaluation of cytokine/chemokine profiles suggests that an active T-cell response is induced during acute infection, even in the absence of seroconversion, and returns to normal levels after treatment and symptom resolution (23). Thus, interrogating the T-cell response may be a useful strategy for detecting LD during the earliest stages of illness and understanding the immune response throughout disease progression.

T-cell responses rely on the capacity of unique T-cell receptors (TCRs) to recognize specific peptide antigens presented on the cell surface by human leukocyte antigen (HLA) proteins for antigen-induced clonal expansion and differentiation into effector cells. Some of these expanded T cells become part of the memory compartment, where they can reside for many years as clonal populations of cells with identical TCR rearrangements (24–26). While the diversity of TCR recombination means that most of these disease-specific TCRs are “private,” or highly unique to one individual, part of the T-cell response to a disease is “public,” with identical amino acid sequences observed across multiple individuals, particularly those with shared HLA backgrounds (27). The public, disease-associated TCRs can be identified using a case/control study design (28–31) and matched to specific antigens through multiplex identification of antigen-specific TCRs (MIRA) (28, 32). Because these public clones are antigen- and HLA-specific, enrichment of such clones serves as a signature of infection in a given HLA context (28, 29). This approach has been successful in identification of past infection by cytomegalovirus (CMV) (28) and severe acute respiratory syndrome coronavirus 2 (SARS-CoV-2) (29, 30, 33).

Given the well-characterized roles of CD8+ and CD4+ T cells in viral control and clearance (34), it is not surprising that strong T-cell responses are elicited by SARS-CoV-2 and mediated by similar antigens and TCRs across individuals (29–31). While CD4+ T cells are known to differentiate into effector cell types with roles in promoting cellular and humoral immunity following many bacterial infections (35), the publicity of T-cell responses to bacterial pathogens has yet to be characterized. To better understand the utility of TCR immunosequencing in the context of bacterial infection, we leveraged the previously described case-control study design approach (28–31) to characterize LD-specific TCRs and develop a classifier to aid in diagnosis of LD. We tested the performance of the classifier in case and control cohorts distinct from those used in classifier training and assessed the correlation between T-cell responses and clinical features of LD. We also mapped a subset of the identified LD-associated TCRs to specific *Bb* antigens, supporting the biologic specificity of the TCR immunosequencing approach.

## RESULTS

### Identification of LD-associated TCRs

To characterize the T-cell response to LD, we immunosequenced TCRβ in samples from 3 LD cohorts and a database of controls from Lyme-endemic and non-endemic regions (Tables 1 and 2). Public TCRs associated with early LD were identified from a subset of these samples comprising a case/control training dataset of 72 patients identified from the LDB (n=54) and Boca (n=18) cohorts who presented with STTT-positive early LD prior to 2019 and control repertoires (n=2,981) from a database of healthy individuals from non-endemic regions recruited for other studies and presumed to be LD-negative (Figure 1A). A total of 251 public TCRs associated with early LD, referred to as “enhanced sequences” (ESs), were identified based on statistical enrichment in cases compared with controls.

**Figure 1.**
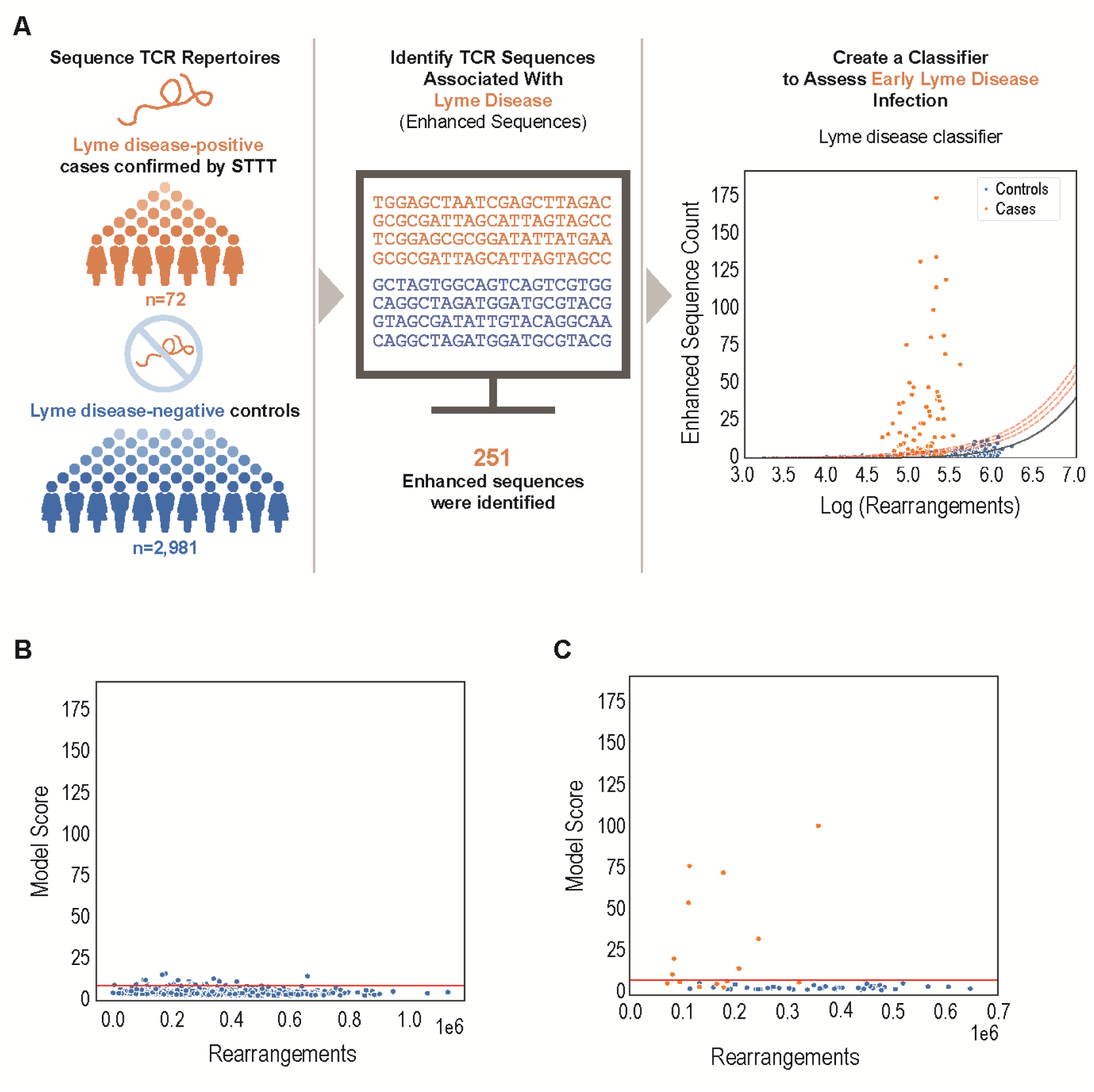
LD-associated TCRs distinguish cases from controls in training cohorts. (A) Development and training of the classifier used to assess *Bb* infection using a T-cell receptor assay. The classifier was trained using TCR repertoires sequenced from 72 STTT–confirmed cases (orange) and 2,981 controls (blue) (see Table 2). A total of 251 LD-associated TCR sequences (enhanced sequences) were identified. Distribution of the number of TCR rearrangements encoding enhanced sequences as a function of the (log-transformed) total number of unique TCR rearrangements identified in a repertoire. Distribution of enhanced sequences in control samples approximately follows a logistic-growth curve (solid black line; dashed red lines indicate +2, +3, and +4 standard deviations from fit), which was used to define a scoring function. (B) Distribution of the model scores is largely invariant to the number of unique rearrangements in an independent set of controls from Lyme-endemic areas (n=2,627). Red line indicates 99th percentile distribution in this cohort (score=4.2675), which was defined as the positive-call threshold. (C) Model score distribution in a holdout set of repertoires from the LDB cohort (n=15 cases; n=48 controls), collected in 2019 and immunosequenced after model training.

**Table 1.** Description and criteria for cohorts used in study.

| Name of Cohort | Cohort | Description/Criteria |
| --- | --- | --- |
| LDB (pre-2019) | Case | Patients presenting prior to 2019:<br>With EM >5 cm in diameter or an erythematous, annular, expanding skin lesion ≤5 cm, diagnosed by a health care provider at study enrollment or<br>With no history of chronic fatigue syndrome, rheumatologic disease, or multiple sclerosis presenting with signs or symptoms (headache, fatigue, fever, chills, or joint or muscular pain) without an EM/annular lesion, but with a suspected tick exposure or tick bite<br>STTT-positive |
|  | Control (endemic) | Healthy individuals enrolled prior to 2019 living in an area of endemicity with no history of Lyme disease or tick-borne infection STTT-negative |
|  | Cross-reactivity case | Symptomatic patients<br>STTT-negative<br>PCR-positive for babesiosis (n=5) or anaplasmosis (n=4) |
| Boca Biolistics | Case | Patients with an EM rash, positive serology results, and/or evidence of a tick bite<br>STTT-positive |
| JHU | Case | Patients presenting with EM ≥5 cm diagnosed by a health care provider at study enrollment<br>Had received ≤72 hours of appropriate antibiotic treatment for early Lyme disease at enrollment |
|  | Control (endemic) | No clinical or serologic history of Lyme disease<br>STTT-negative |
| LDB (2019) | Case | Patients presenting during 2019 with the same characteristics described for the LDB case subgroup, except positives were confirmed by STTT and/or PCR |
|  | Control (endemic) | Healthy individuals enrolled during 2019 with the same characteristics described for the LDB controls from Lyme-endemic areas subgroup |
| Database | Control (endemic) | Individuals with unknown Lyme disease status living in Lyme-endemic regions in the United States and Europe |
| Database | Control (non-endemic) | Individuals with unknown Lyme disease status living in Lyme non-endemic regions in the United States |
| DLS cohort | Cross-reactive | Suspected *Babesia* samples (PCR-positive) |

**Table 2.** Cohorts used for training, setting the classification threshold, verification and determination of sensitivity and specificity of the classifier.

| Study Phase | Name of Cohort | Lyme Disease Status | Samples Used |
| --- | --- | --- | --- |
| Classifier training | LBD (pre-2019) | Case | 54 |
|  | Boca Biolistics | Case | 18 |
|  | In-house database | Control (non-endemic) | 2,981 |
| Setting threshold | In-house database | Control (endemic) | 2,507 |
|  | LDB (pre-2019) | Control (endemic) | 120 |
| Verification (holdout set) | LDB (2019) | Case | 15 |
|  | LDB (2019) | Control (endemic) | 48 |
| Sensitivity | JHU | Case | 211 |
| Specificity | LDB (pre-2019) | Control (endemic) | 115 |
|  | In-house database | Control (endemic) | 2,471 |
|  | JHU | Control (endemic) | 45 |
|  | DLS | Control (suspected anaplasmosis) | 12 |
| Cross-reactivity | LDB (pre-2019) | Cross-reactivity (endemic) STTT-negative, PCR-confirmed cases of anaplasmosis and babesiosis | 9 |

### Enhanced TCR sequences are highly specific for identifying early LD

Clonal expansion in response to *Bb* infection should lead to enrichment of LD-associated ESs in patients with LD. A low-level presence of LD-associated ESs would also be expected among healthy individuals. The observed background rate of ESs is a function of the number of sequenced T cells and the number of unique TCRs in each sample. Empirically, the background number of ESs in our training population fit a logistic-growth curve. To leverage the number of ESs and total unique productive TCR rearrangements as a diagnostic classifier, we modeled the number of ESs as a logistic-growth function of the number of unique productive TCRs sampled from a repertoire and fit this model to the 2,981 control repertoires in the training data (Figure 1A, right graph; black line represents the model fit). The resulting model provides a normalized estimate of the degree to which the ES signature of a case sample deviates from what is typically seen in our control data, expressed in standard deviations from the mean (Figure 1A, right graph, red dashed lines). This approach carefully controls specificity by considering thousands of control repertoires. The final positive/negative call threshold was set to a specificity of 99% on an independent set of controls from Lyme-endemic areas (n=2,627, consisting of 2,507 presumed LD-negative samples from our database and 120 STTT-negative controls from Lyme-endemic areas from the LDB cohort [Figure 1B; Tables 1 and 2]).

To further verify the classifier and call threshold among patients with demographics similar to the training dataset, we applied the resulting model to a holdout set of samples from the LDB cohort collected in 2019 that included 15 laboratory-confirmed–positive (by STTT and/or polymerase chain reaction [PCR]) cases of early LD and 48 laboratory-confirmed–negative (by STTT) controls from Lyme-endemic areas with no history of Lyme or tick-borne infection (Figure 1C; Tables 1 and 2). Overall, 8 of 15 (53%) early LD samples and 0 of 48 (0%) controls from Lyme-endemic samples were identified as TCR-positive by the classifier.

### Sensitivity of TCR repertoire analysis identifying early LD

To further assess the generalizability of our approach in an independent cohort collected with different protocols, we evaluated performance of the TCR classifier using samples from STTT- positive and STTT-negative patients who had clinically diagnosed early LD and documented EM at the time of their enrollment in the JHU cohort (Tables 1 and 2). Consistent with results from the LDB and Boca cohorts, 118 of 211 (56%) patients diagnosed with early LD were classified as TCR-positive (Table 3), while only 32 of 2,631 (1.2%) control samples from Lyme-endemic areas were TCR-positive (0/115 from LDB, 1/45 from JHU, and 31/2,471 from a database of individuals with unknown LD status living in Lyme-endemic regions in the United States and Europe) (Figure 2A). In contrast, only 64 of 211 (30%) LD-positive JHU samples were STTT- positive, representing a 1.9-fold reduction in sensitivity relative to TCR immunosequencing. To further confirm the specificity of the TCR signature, we evaluated 21 samples from individuals with PCR-confirmed anaplasmosis (n=4) or babesiosis (n=17) from the LDB and DLS cohorts. All 21 samples were negative according to the classifier (Figure 2A).

**Figure 2.**
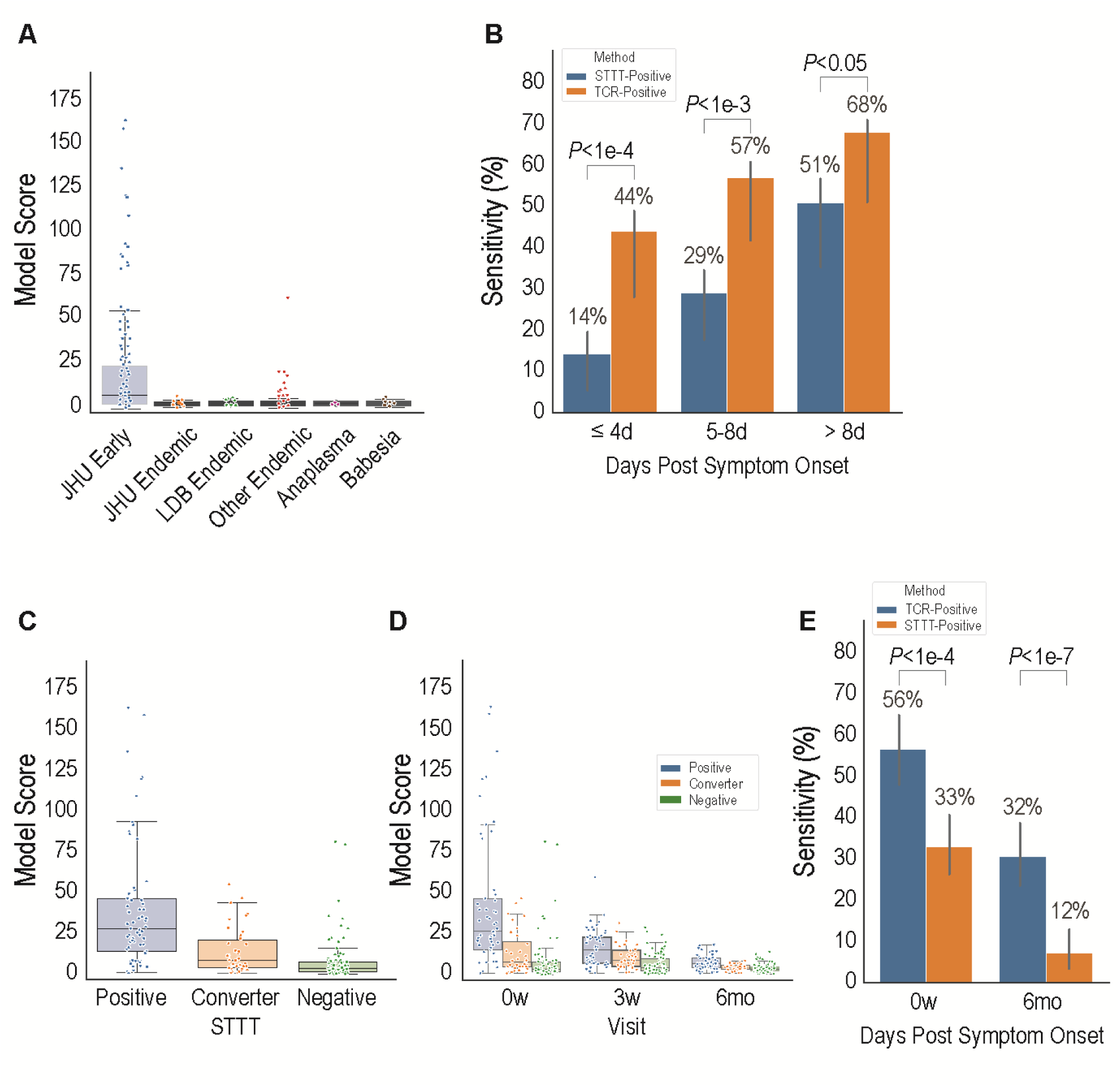
Validation of the TCR classifier in the JHU cohort and other holdout endemic controls. (**A**) Model score distribution in early LD samples from JHU (blue, n=211), in addition to holdout controls from Lyme-endemic areas. JHU (orange, n=45) and LDB (green, n=115) controls from Lyme-endemic areas were LD-negative based on clinical assessment and negative STTT testing. Other controls from Lyme-endemic areas (red, n=2,471) were drawn from our database of repertoires sampled from individuals in endemic regions in the United States and Europe who were presumed negative for LD. The *Anaplasma* samples (purple, n=4) were from the LDB cohort, and the *Babesia* samples were from the LDB and DLS cohorts (brown, n=5 and n=12, respectively). (**B**) Sensitivity of STTT and the TCR classifier for individuals in the JHU cohort, stratified by symptom duration (days) at time of enrollment, along with bootstrapped 95% CI. Participants were stratified based on self-reported symptom duration into bins of <u>≤</u>4 days (n=73), 5-8 days (n=75) or >8 days (n=63). The sensitivities of both TCR testing and STTT increased with longer duration of symptoms reported at the time of testing. (**C**) Model score distribution for JHU early disease samples stratified by STTT serostatus at enrollment and posttreatment follow-up. Positive (blue, n=64): STTT-positive at enrollment; converter STTT (orange, n=38): STTT-negative at enrollment and STTT-positive at posttreatment follow-up; negative (green, n=109): STTT-negative at both visits. (**D**) Longitudinal dynamics of TCR scoring by serostatus for the JHU cohort. Positive (blue, n=53): STTT-positive at enrollment; converter (orange, n=32): STTT-negative at enrollment and STTT positive at posttreatment follow-up; negative (green, n=76): STTT-negative at both visits. **(E)** For the 163 patients who had STTT performed pretreatment and at 6-month follow-up, difference in sensitivities was calculated for recent Lyme infection between STTT and TCR test. In box-and-whisker plots, boxes indicate median ± IQR, and whiskers denote 1.5 times the IQR above the high quartile and below the low quartile. Significant differences in sensitivity were evaluated by T test.

**Table 3.**
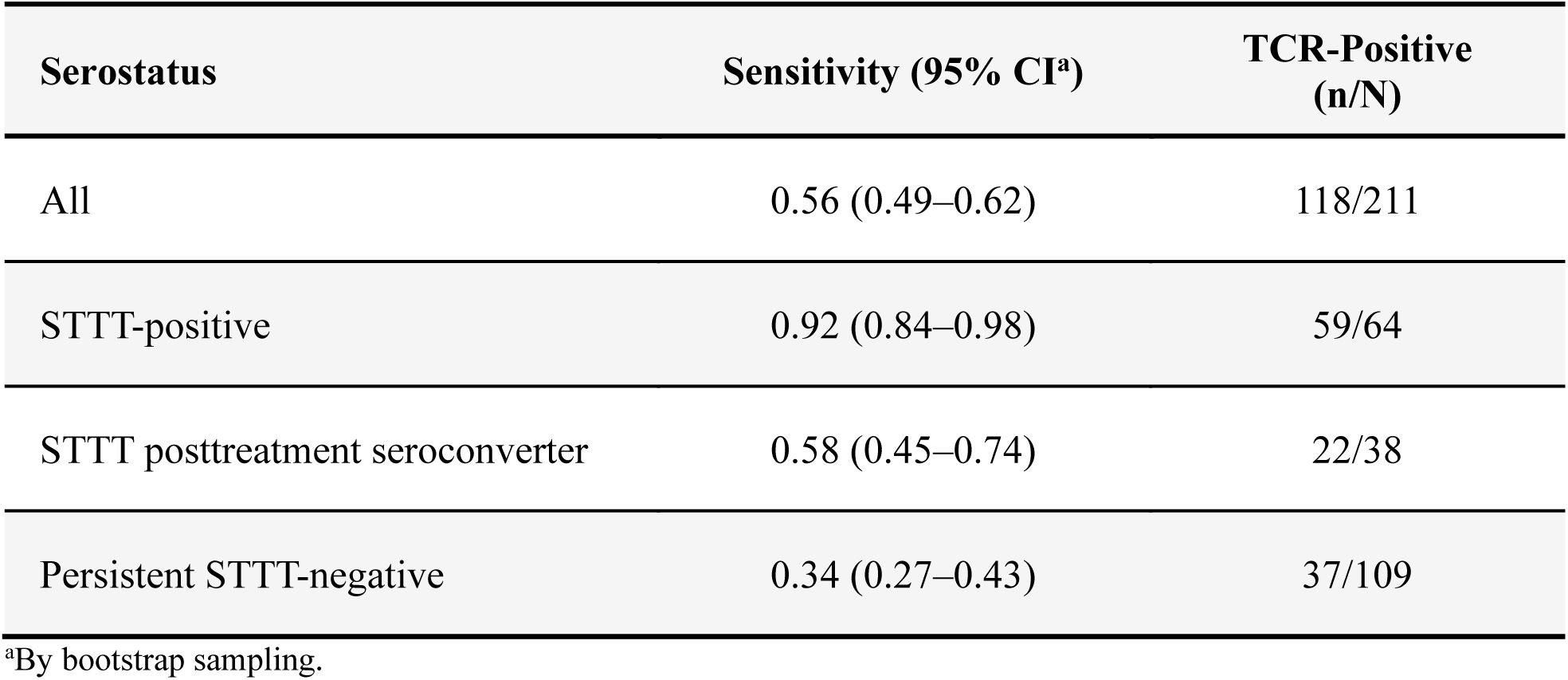
TCR classifier sensitivity stratified by serostatus in the JHU cohort.

| Serostatus | Sensitivity (95% CI<sup>a</sup>) | TCR-Positive (n/N) |
| --- | --- | --- |
| All | 0.56 (0.49–0.62) | 118/211 |
| STTT-positive | 0.92 (0.84–0.98) | 59/64 |
| STTT posttreatment seroconverter | 0.58 (0.45–0.74) | 22/38 |
| Persistent STTT-negative | 0.34 (0.27–0.43) | 37/109 |

<sup>a</sup>By bootstrap sampling.

When examined as a function of self-reported duration of symptoms, sensitivity of T-cell and antibody tests increased with time, yet T-cell testing showed greater sensitivity at early time points (Figure 2B). If T-cell responses typically precede and facilitate B-cell responses in response to *Bb* infection, consistent with immune response to viral and bacterial pathogens (36, 37), then TCR positivity among a seronegative cohort should predict subsequent seroconversion, and a majority of STTT-positive samples should also be TCR-positive. Indeed, among the JHU case cohort, TCR testing was positive in 59 of 64 (92%) STTT-positive cases (58 of 61 [95%] STTT-positive by IgM), compared with 59 of 147 (40%) STTT-negative cases (Table 3), indicating that a detectible antibody response is highly predictive of a detectible T-cell response. Importantly, of the 59 TCR-positive/STTT-negative individuals, 22 (37%) subsequently seroconverted from STTT-negative at study enrollment to STTT-positive at the first post-treatment follow-up visit (∼3 weeks after enrollment), while only 16 of 88 (18%) individuals who were TCR-negative at baseline seroconverted over the same time period (*P*=0.01, Fisher’s exact test). Stratification of the JHU cohort demonstrated that median TCR model scores (Figure 2C) and classifier sensitivity (Figure S3 Table 3) were highest among individuals who were STTT-positive at enrollment, intermediate among those who seroconverted post treatment, and lowest among individuals who remained persistently STTT-negative, highlighting differences in the extent of LD-associated T-cell expansion in these serologically defined subpopulations.

Taken together, these data indicate that detectable LD-associated T cells typically expand prior to detectible antibodies, suggesting that identification of LD may be aided by TCR immunosequencing during early phases of *Bb* infection.

### Disease-associated TCRs and seropositivity wane after treatment

Previous data suggest that T-cell and humoral immune responses exhibit differing dynamics over the course of *Bb* infection (21). To probe these dynamics, we evaluated TCR repertoires in longitudinal samples from individuals enrolled in the JHU cohort. Patients were either antibiotic-naïve or had initiated 3 weeks of oral doxycycline treatment within the 72 hours prior to study enrollment, with samples collected at enrollment, immediately after treatment, and at 6 months post treatment. Immunosequencing of samples from 161 patients with samples available at all time points revealed that TCR responses waned significantly in the 6 months following treatment (Figure 2D) (30). Median model scores decreased from 6.1 to 2.5, and model sensitivity decreased from 56% (91/161) at enrollment to 32% (51/161) 6 months post treatment. Notably, the sensitivity of STTT also declined over the same time period, from 33% at enrollment to 12% at 6 months post treatment (14 of 115 patients with available STTT results) (Figure 2E). Similar to the results shown in Figure 2C, TCR model scores were higher across all time points among individuals who were STTT-positive at baseline compared with those who were STTT-negative (Figure 2D). T-cell testing was more sensitive than STTT for identification of LD post treatment (32% vs 12% at 6 months post treatment (Figure 2E) including in patients who did not undergo IgG seroconversion (not shown).

### T-cell responses correlate with clinical measures of LD severity

The strong correlation observed between antibody and T-cell responses highlights the interconnectedness of the immune response in early LD and may also reflect underlying pathogen burden, disease severity, or other clinical measures that drive the immune response. We therefore explored potential associations between clinical parameters previously reported in the JHU study (38) and the strength of the T-cell response as measured by the TCR model score at diagnosis. In both univariate analyses (Figure 3A-D) and a multiple regression model (Table S1) that adjusted for sex, age, and serostatus, higher TCR scores were associated with markers of disease severity, including ≥1 elevated liver function test (aspartate aminotransferase, alanine aminotransferase, and alkaline phosphatase), disseminated rash, and the number of pretreatment LD-associated symptoms. Notably, the highest model scores were observed among STTT- positive individuals with disseminated rash and elevated liver function tests (Figure 3A, 3C, S2).

**Figure 3.**
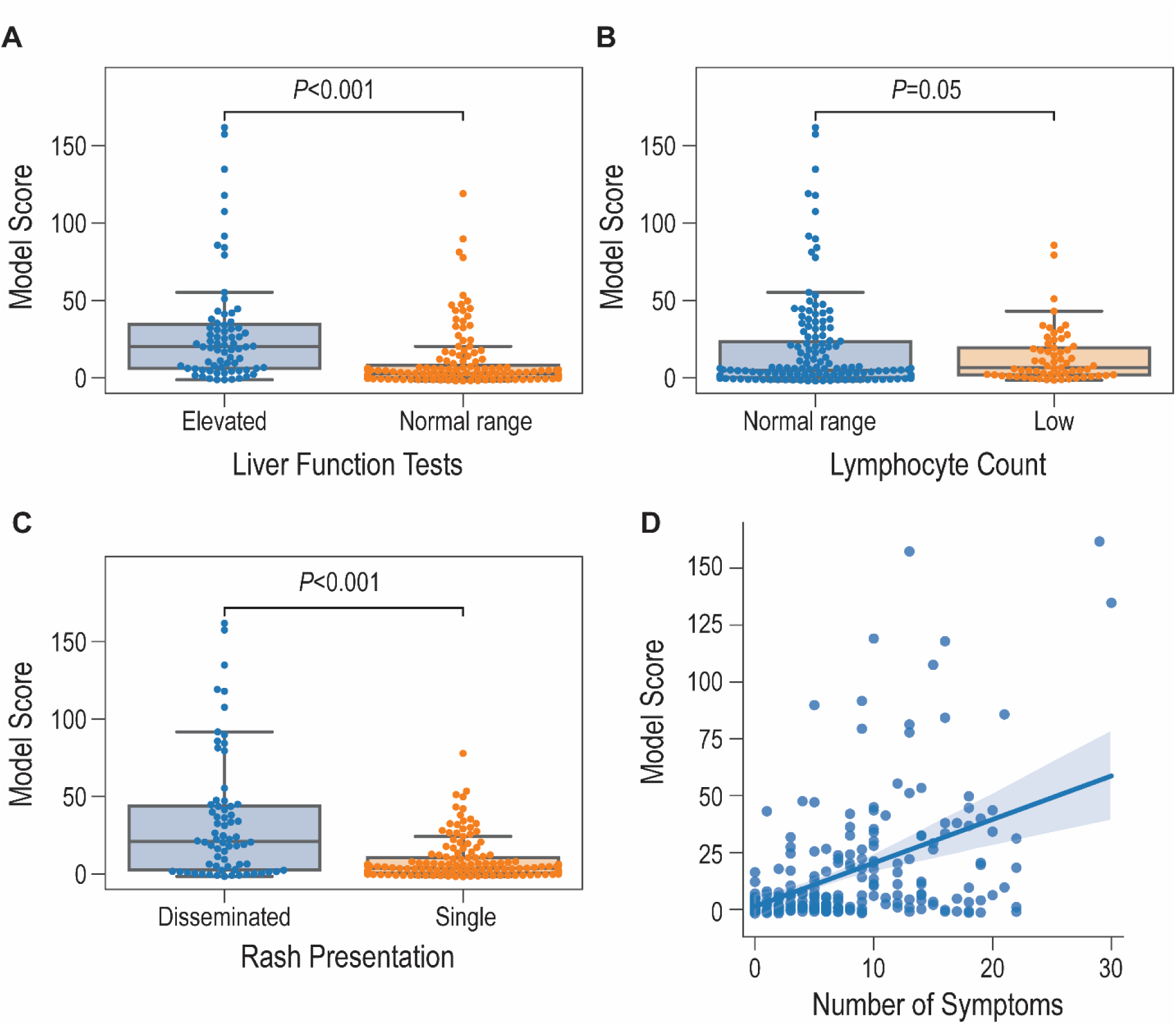
Clinical correlates of TCR scoring. TCR scores were stratified by (**A**) liver function test results (elevated [n=72] vs normal [n=139]), (**B**) lymphocyte counts (normal [n=150] vs low [n=61]), or (**C**) presentation of rash (multiple/disseminated [n=68] vs single [n=143]) and (**D**) plotted as a function of the number of Lyme-related symptoms (Spearman R^2^=0.17. *P* values, Mann-Whitney *U* test). In box-and-whisker plots, boxes indicate median ± IQR, and whiskers denote 1.5 times the IQR above the high quartile and below the low quartile.

Sex, age, size of rash, and an abnormally low lymphocyte count were not associated with a difference in TCR model scores in this cohort (Table S1).

### Antigen specificity of Lyme-associated TCRs

Analysis of TCR sequence similarity clustered 105 of the 251 ESs (42%) to one of 6 clusters, 5 of which were statistically associated with an HLA class-II heterodimer (Table 4 and Supplementary Materials). In a MIRA experiment that queried T cells derived from 395 healthy donors against 777 peptides from 26 *Bb* proteins, 6 ESs from *HLA-DRB3*02:02*–associated cluster 6 exactly matched TCRs that were mapped to the flagellin B (FlaB)-derived peptide MIINHNTSAINASRNNG, providing direct evidence of *Bb* specificity for these ESs. One of these TCR sequences was found in 26 individuals, all of whom expressed *HLA-DRB3*02:02* (among the 25 with available typing). A Basic Local Alignment Search Tool (BLAST) analysis of this peptide showed limited sequence similarity with non-*Borrelia* pathogens, and the associated TCRs were highly enriched in JHU cases compared to endemic controls (Fig. 4).

**Figure 4.**
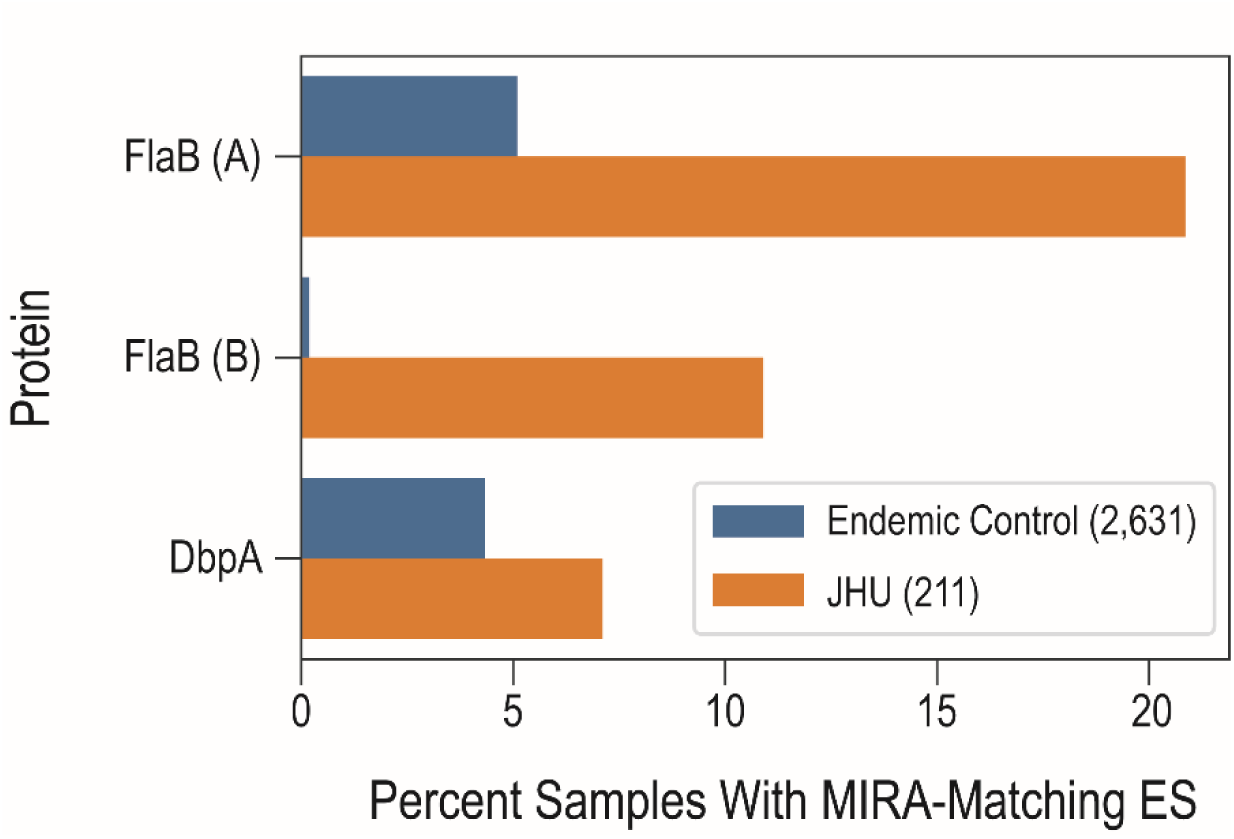
Percentage of early LD samples with ESs assigned by MIRA to the indicated *Bb* antigens. FlaB (A/B), flagellin protein B antigen A/B; DbpA, decorin-binding protein A.

**Table 4.**
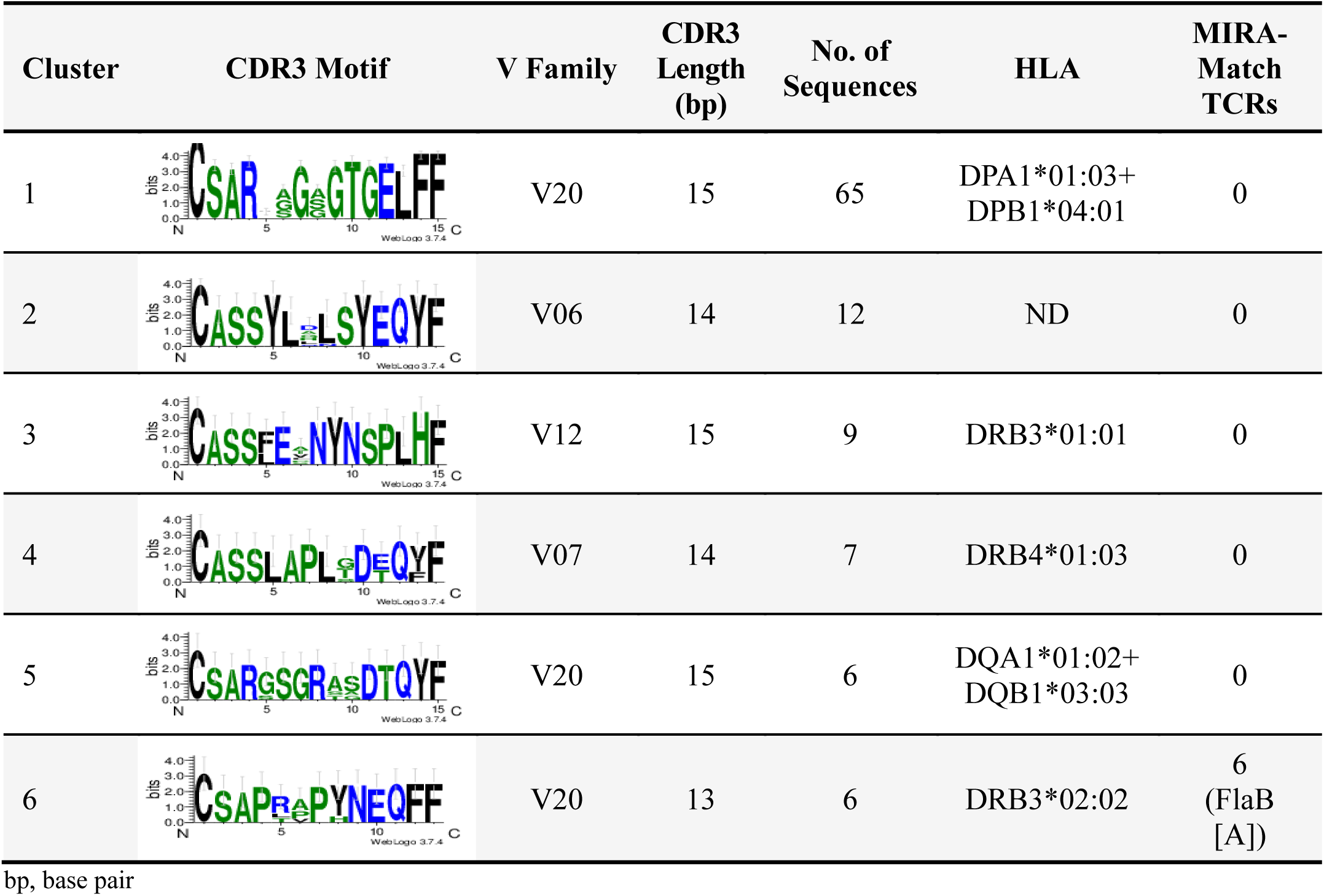
TCR clusters as defined by connected components within a 1–amino-acid change in CDR3 and the same V-gene family.

| Cluster | CDR3 Motif | V Family | CDR3 Length (bp) | No. of Sequences | HLA | MIRA-Match TCRs |
| --- | --- | --- | --- | --- | --- | --- |
| 1 |  | V20 | 15 | 65 | DPA1*01:03+<br>DPB1*04:01 | 0 |
| 2 |  | V06 | 14 | 12 | ND | 0 |
| 3 |  | V12 | 15 | 9 | DRB3*01:01 | 0 |
| 4 |  | V07 | 14 | 7 | DRB4*01:03 | 0 |
| 5 |  | V20 | 15 | 6 | DQA1*01:02+<br>DQB1*03:03 | 0 |
| 6 |  | V20 | 13 | 6 | DRB3*02:02 | 6<br>(FlaB [A]) |

bp, base pair

Three additional non-clustered TCRs were mapped to 3 separate antigens, 2 in FlaB and 1 in Dbpa, and each of these were similarly enriched in cases compared to controls (Fig. 4). As a negative statistical control, none of the LD ESs matched any TCRs derived from 507 individuals that were previously queried against 325 SARS-CoV-2–derived peptides.

## DISCUSSION

Results from this study provide proof of principle that the high-throughput TCR sequencing and machine learning approach we previously applied for identification of viral infections (eg, CMV and SARS-CoV-2 [28, 29, 30]) can be adapted for identification of *Bb* infection. We demonstrate that TCR repertoire characterization for sequence-based identification of public, disease-specific TCRs in the setting of acute bacterial infection is a powerful and generalizable approach to aid in diagnosing disease.

Identification of 251 LD-associated enhanced TCR sequences served as the basis for training a classifier capable of sensitive and specific detection of LD across 3 independent cohorts of patients with laboratory-confirmed and/or clinically diagnosed early LD. The TCR classifier identified patients with early LD with 1.9-fold greater sensitivity than STTT (56% vs 30%), while maintaining a specificity of 99%. Enhanced sensitivity was most apparent in early illness (44% vs 14%, or 3.1-fold increase in sensitivity ≤4 days since symptom onset), and TCR positivity was predictive of subsequent STTT seroconversion in 37% of STTT-negative individuals, consistent with expansion of LD-specific T cells preceding detectable antibody responses.

TCR positivity was associated with STTT positivity at enrollment (92%) and did not decline as rapidly as serologic responses following treatment. Longitudinal analyses showed that TCR scores decreased with time post treatment, consistent with diminishment of the T-cell response with resolution of disease, yet remained more sensitive than STTT for identification of LD post treatment, including in patients who did not undergo IgG seroconversion. This observation suggests that TCR testing may be able to identify LD even in the absence of seroconversion at convalescence, which has been observed among individuals treated early in the course of disease (39). Furthermore, higher TCR scores correlated with clinical measures of disease, including elevated liver function tests, disseminated rash, and number of disease-associated symptoms. This suggests that the magnitude of the T-cell response is associated with the degree of disease severity. Finally, we mapped a subset of Lyme-associated ESs to known *Bb* antigens, supporting the high biologic specificity of a TCR immunosequencing approach (40).

Our data imply that T-cell activation precedes the humoral response, although both T-cell‒ dependent and –independent responses have been implicated in clearance of *Borrelia* infection (37, 41). Currently, the primary CDC-recommended testing strategy for LD is STTT, which probes the humoral response to *Bb* (7, 8). Given the high prevalence of testing performed in patients during the early stages of LD infection, when sensitivity of STTT is poor (9), alternative approaches to detecting *Bb* are needed. Recently, modified two-tiered testing (MTTT) algorithms have been adopted that utilize 2 sequential ELISAs, eliminate immunoblotting, and demonstrate improved sensitivity over STTT in early LD (42). Even so, almost half of individuals with PCR- confirmed *Bb* infection do not produce a detectable serologic response (43). Our results confirm the ability of the TCR assay to identify LD in a large proportion of STTT-negative individuals prior to seroconversion. These data support further studies directly comparing TCR testing to MTTT to better understand the potential of TCR testing as both an alternative and complementary diagnostic approach to any serologic testing modality.

The present analysis is limited to retrospective evaluation of samples previously collected from well-defined, prospective cohorts of clinically confirmed and/or laboratory-confirmed early LD. Additional prospective clinical validation studies are needed to further characterize the advantages of TCR testing relative to serology in scenarios where the spectrum of presenting illness, symptomology, and duration may vary. For example, further studies are needed to understand the utility of TCR testing in patients with persistent symptoms in the setting of LD. Approximately 10% to 20% of patients treated for LD experience long-term symptoms lasting 6 months or more after treatment, known as post-treatment Lyme disease (PTLD) (6, 44). TCR testing may provide a means for diagnosing these patients and minimize the potential for misleading interpretation of positive results (23, 45). In addition, we found that 11 of 12 individuals who presented with EM rash within the JHU cohort, but were both STTT-negative and *Bb* PCR-negative, were also TCR-negative. These data may reflect sensitivity limitations of both immune assays but could also be attributable to difficulties in clinically discriminating between LD and other similar tick-borne illnesses, such as STARI (43), suggesting exploration of TCR testing for differential diagnosis of LD and other similar tick-borne illnesses as another important area of future investigation. Although we examined potential assay cross-reactivity against *Anaplasma* and *Babesia* (Figure 2A), evaluation of cross-reactivity with other pathogens should be performed. Nevertheless, it is noteworthy that antigen mapping indicates that a subset of the identified TCRs are highly specific for known *Bb* proteins, including FlaB, a protein known to bind cross-reactive antibodies in the immunoblotting component of STTT (46).

Results of this study suggest that TCR testing can have high clinical utility as a sensitive and specific diagnostic for LD that may facilitate earlier diagnosis of LD and initiation of antibiotic treatment to prevent development of severe illness in patients lacking definitive clinical signs/symptoms. Application of the present TCR classifier as a diagnostic assay will be prospectively evaluated relative to 2-tiered testing in an ongoing clinical validation study in patients presenting with suspected LD (NCT04422314).

## MATERIALS AND METHODS

### Study design

This study employed previously collected samples to develop and validate a model for identifying early LD with high sensitivity and specificity. TCR repertoires were sequenced from 8,585 participants, including 298 cases of LD enrolled in prospective studies and 8,287 controls obtained from a variety of sources. Allocation of samples to sets used for classifier training, setting the classification threshold, and validation of the TCR assay was prespecified; all available samples meeting the quality control (QC) and assignment criteria were included in the analysis (see Assignment of Cohorts and Tables 1 and 2). The primary endpoint of the study was evaluation of sensitivity in the JHU cohort, which was selected based on the conservative enrollment criteria for that cohort. Inclusion and exclusion criteria for all cohorts are detailed below.

All procedures involving human participants were conducted in accordance with the ethical standards of the 1964 Declaration of Helsinki and its later amendments or comparable ethical standards. For the LDB cohort, institutional review board (IRB) approval was obtained for each site through the LDB sponsor protocol (Advarra IRB) or the institution-specific IRB. For the Boca cohort, IRB approval was obtained through the Advarra IRB. Human subject protocols for the JHU cohort were approved by the IRBs of Johns Hopkins University and Stanford University. All participants provided written informed consent prior to enrollment.

### Study cohorts

For a detailed summary of all cohorts, see Tables 1 and 2.

#### LDB cohort

LDB is a program of the Bay Area Lyme Foundation. The LDB cohort enrolled individuals from the East coast and upper Midwest regions of the United States who presented with signs or symptoms consistent with early LD. Included patients presented with EM rash greater than 5 cm in diameter or an erythematous, annular, expanding skin lesion of 5 cm or less or presented with signs or symptoms (headache, fatigue, fever, chills, or joint or muscular pain) without an EM/annular lesion, but with a suspected tick exposure or tick bite, and with no history of chronic fatigue syndrome, rheumatologic disease, or multiple sclerosis. Individuals with tick-bite reactions (eg, a non-annular erythematous macule at the site of the tick bite) without EM or expanding annular lesion were excluded, as were those who had initiated antibiotics more than 48 hours before enrollment. Healthy individuals living in the same regions with no history of LD or tick-borne infection were enrolled as controls from Lyme-endemic areas. Real-time PCR for *Bb* detection in whole blood and serologic testing for antibodies against *Bb* using the immunoassays comprising STTT (non-reflexive) were conducted for all individuals.

Laboratory-confirmed early LD samples were defined as being STTT-positive, PCR-positive (sample or culture), or having 2 positive ELISAs and an EM of 5 cm or larger in diameter.

Laboratory-confirmed controls from Lyme-endemic areas were defined as STTT-negative. Additional details and baseline clinical characteristics of this cohort were previously published (47). Only laboratory-confirmed samples that were STTT-positive were used in model training.

Only laboratory-confirmed–positive (by STTT and/or PCR) cases and laboratory-confirmed– negative (by STTT) controls from Lyme-endemic areas were included in analyses of model performance or to establish the final call threshold. Nine cross-reactivity case samples that were positive by PCR for *Anaplasma* (n=4) or *Babesia* (n=5) were used for additional specificity testing.

#### Discovery Life Sciences (DLS) cohort

Blood samples from 12 individuals found to be *Babesia*-positive by PCR testing were acquired from Discovery Life Sciences for use in specificity testing.

#### Boca cohort

Specimens were collected from antibiotic-treatment–naïve patients recruited at clinical sites throughout New York and New Jersey who presented with acute symptomology of a tick-borne illness. Participants had blood drawn on 3 occasions: 30 days or less post-tick bite while antibiotic-treatment–naive, 6 to 8 weeks post tick bite, and 16 to 24 weeks post tick bite. Whole blood samples were aliquoted, frozen, and stored at −80°C after collection. At each visit, information was captured regarding symptoms, date of symptom onset, treatment status, treatment regimen, and lab results. Specimens were received at Boca Biolistics Reference Laboratory (Pompano Beach, FL, USA) and characterized for relevant tick-borne pathogens, including *Bb*, *Babesia microti*, *Ehrlichia chaffeensis*, and *Anaplasma phagocytophilum*. Testing was performed in-house and at ARUP Laboratories (Salt Lake City, UT, USA) on matched serum collected from donors. DiaSorin and Immunonetics Lyme antibody testing was performed at Boca Biolistics Reference Laboratory, and IgM- and IgG-specific antibody screening for *Bb*, *B. microti*, *E. chaffeensis*, and *A. phagocytophilum* was performed at ARUP Laboratories. For the present study, immunosequencing was performed on the first available sample from 18 donors who were seropositive for *Bb* by ELISA and immunoblot (either IgG or IgM), all of whom were classified as STTT-positive by 2-tiered testing criteria.

#### JHU cohort

The SLICE study is a longitudinal, prospective cohort study that enrolled patients 18 years of age or older with early LD who were self-referred or recruited from primary or urgent care settings from 2008 to 2020. Eligible participants were enrolled primarily at study sites in Maryland, with a small number enrolled at a satellite site in Southeastern Pennsylvania. At enrollment, participants were required to have a visible EM of at least 5 cm in diameter diagnosed by a healthcare provider. All patients had received no more than 72 hours of appropriate antibiotic treatment for early LD at enrollment. Additional details and baseline clinical characteristics have been previously published (48). Participants without a clinical or serologic history of LD were recruited from similar primary care settings or through the community using flyers and online advertising to serve as controls from Lyme-endemic areas. Controls were required to be STTT- negative at the time of enrollment and at all subsequent visits. Participants in both groups were excluded for a range of self-reported prior medical conditions paralleling those listed in the proposed case definition for post-treatment LD syndrome (49), specifically, chronic fatigue syndrome, fibromyalgia, unexplained chronic pain, sleep apnea or narcolepsy, autoimmune disease, chronic neurologic disease, liver disease, hepatitis, HIV, cancer or malignancy in the past 2 years, major psychiatric illness, or drug or alcohol abuse.

Patients with early LD were treated with 3 weeks of oral doxycycline (5) and seen regularly over the course of 1 to 2 years. Samples collected before treatment, immediately after treatment, and 6 months post treatment were used for the present study, in addition to samples collected from control participants at an initial study visit. Disseminated EM rash was defined as more than 1 rash site visible on physical exam at the pretreatment study visit, and local rash was defined as a single EM rash site. Duration of illness was determined by self-report of the number of days since the first appearance of LD-specific signs or symptoms. The number of LD symptoms at enrollment was obtained through an interviewer-administered questionnaire. STTT status was determined by Quest Diagnostics at each study time point using CDC recommendations incorporating duration of illness at time of testing (7).

High-resolution HLA class I/II typing for the JHU cohort (cases only) was performed by Scisco Genetics, Inc., (Seattle, WA, USA) using the ScisGo HLA v6 typing kit, as previously described (50, 51).

#### Database controls

A total of 7,959 repertoires sampled during previous studies were selected from our database. Inclusion was determined at the cohort level and based on the size of the cohort, geographic region (United States and LD-endemic regions of Europe), and sequencing date (2019 or later, to ensure consistent lab sequencing protocols). Samples were classified as being either from endemic regions (Germany, Italy, or upper Midwest or Northeast regions of the United States) or non-endemic regions (other regions of the United States). All individuals in these cohorts were not tested but were presumed to be LD-negative.

### Assignment of cohorts

Training cases were drawn from the LDB and Boca cohorts (Tables 1 and 2). To enrich for cases with a likely immune response and maximize our ability to detect LD-associated ESs, the training set was limited to 72 STTT-positive cases (54 LDB, 18 Boca). Training controls included 2,981 repertoires from individuals from non-endemic regions of the United States and Europe previously collected as part of other studies and presumed to be LD-negative.

The positive-call threshold was set based on 2,507 presumed LD-negative samples collected from endemic regions available in our database, along with 120 confirmed STTT-negative controls from Lyme-endemic areas randomly selected from the LDB cohort. Additional LDB case (n=15) and control (n=48) samples collected during the 2019 tick season were sequenced after model training and used as an initial check of model specificity and generalizability.

TCR assay sensitivity was evaluated in the JHU cohort. Repertoires sampled from 211 participants at the time of enrollment passed QC thresholds established after model training described below. A subset of patients in the JHU cohort (n=161) had sequenced repertoires that passed QC from samples collected before and after treatment and 6 months post treatment.

Specificity of the final model was estimated based on 3 control cohorts from Lyme-endemic areas: (1) all controls from Lyme-endemic areas from JHU (n=45); (2) 50% (selected by random sampling) of controls from Lyme-endemic areas from tick seasons prior to 2019 in the LDB cohort (n=115 passed QC); and (3) 50% (selected by random sampling) of presumed controls from Lyme-endemic areas from our database (n=2,471 passed QC).

### TCR repertoire immunosequencing

Immunosequencing of TCRβ CDR3 was performed using the immunoSEQ^®^ Assay (Adaptive Biotechnologies, Seattle, WA, USA) and analyzed as previously described (52–54). For additional details, see the Supplementary Materials and Methods.

### Identification of LD-associated ESs

Public TCRβ amino acid sequences associated with early LD were identified as described previously (28). Briefly, one-tailed Fisher’s exact tests (FETs) were performed on all unique TCR sequences to compare frequencies of ESs in early Lyme samples with those in presumed-negative controls. Unique sequences were defined based on the V-gene, J-gene, and CDR3 amino acid sequence. The *P*-value threshold for including a TCR in the ES list was treated as a hyperparameter and was selected to maximize model performance as described below. The resulting set of FET-defined ESs for cohort 𝒞 are denoted *S_fet_^𝒞^*.

A recent study in CMV demonstrated that many TCRs that are not identified as significant ESs in small training datasets based solely on FET may be selected in larger training sets (54). This observation motivates a simple classification problem: prediction of whether a TCR will be identified as an ES by FET when the dataset grows to a specified size. To this end, a logistic regression model was fitted, where the training data were the set of TCRs and corresponding features observed in a previously reported cohort 𝒞_*CMV*_ labeled for CMV serostatus (28). The dependent binary variable was defined as 1 if the TCR was observed as in *S_fet_*^𝒞∗^ for some large CMV-labeled cohort *C*_CMV_^∗^, and 0 otherwise. For each TCR, the following features were defined: (1) average and maximum convergent recombination (CR) for cases and controls; (2) average and maximum productive frequency for cases and controls; and (3) the number of sequences in *S*_*fet*_ similar to the TCR, defined as sharing the same V-gene, having identical CDR3 length, and differing by 1 amino acid.

In practice, a larger CMV-labeled cohort was unavailable. However, as more than 50% of North American and European populations are seropositive for CMV (55), we applied a pseudo- labeling procedure to construct *C*^∗^ . Briefly, a logistic regression classifier (as defined in *[44]*) based on *S*_*fet*_ ^𝒞*CMV*^ was trained on the labeled CMV cohort 𝒞_*CMV*_, then applied to all samples in the LD training cohort. The inferred CMV status was then treated as observed and combined with 𝒞 , resulting in 𝒞^∗^_CMV_ , which was used to define *S*_*fet*_^𝒞*_*CMV*_^. The resulting logistic regression classifier was able to accurately predict which TCRs observed in 𝒞_*CMV*_ but not in *S*_*fet*_^𝒞*CMV*^ would end up in *S*_*fet*_^𝒞*_*CMV*_^ (area under the receiver operating characteristic [ROC] curve=0.84 in cross-validation; data not shown). Features receiving the greatest weight in this model were the CR counts in cases (likely indicating substantial clonal expansion) and the number of similar sequences in *S*^𝒞*CMV*^ (likely indicating that the TCR responds to the same antigen as another ES).

The model fitted to CMV was used to infer ESs for LD. Combining these inferred ESs with ESs identified by FET resulted in the final set of LD-associated ESs, *S^𝒞_Lyme_^*. Hyperparameters in this model were chosen using cross-validation in the context of the disease classification model described below.

### Inferring early LD status based on ES counts in TCR repertoires

Given a set of ESs *S*, the pair (*y*_*i*_, *x*_*i*_) can then be defined for each repertoire *i*, where *x*_*i*_ is the total number of unique productive DNA TCR rearrangements in the sampled repertoire, and *y*_*i*_ < *x*_*i*_ is the number of those rearrangements that encode any of the ESs in *S*. If *y*_*i*_ is treated as sampled from a random variable *Y*, the expected value of *Y* given *x* can be considered. By the way ESs are defined, the distribution of *Y*|*x* is expected to vary substantially between cases and controls.

While this could be treated as a classification problem to maximize the separation between cases and controls (as in [39, 40]), *Y*|*x* was instead explicitly modeled among control samples, with classification based on standard units of deviation above and below expectation. This approach provides superior control of specificity across populations given the extremely unbalanced nature of our case/control dataset. To model this distribution, *Y* was assumed to follow a binomial distribution, with mean *f*(*x*) = *y*_max_ *p*(*x*) and variance *σ*^2^(*x*) = *y*_max_ *p*(*x*)(1 − *p*(*x*)), where *y*_max_ is the maximum number of ESs observed in any training repertoire, and

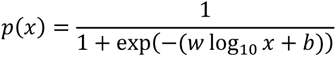

for model parameters *w* and *b*. For a given (*y*_*i*_, *x*_*i*_), the number of standard deviations *y*_*i*_ is from the expected mean given *x*_*i*_ is then used as the model score:

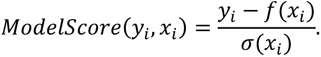

The model parameters *w* and *b* were chosen by minimizing the sum of squared residuals over the set of training control samples.

The observed data are moderately overdispersed with respect to the estimated variance (Figure 1A). As such, the final call threshold *t* was chosen to fix the prespecified false-positive rate of 1% on a set of 2,627 presumed LD-negative control samples as described above.

For a detailed description of TCR repertoire QC criteria, see the Supplementary Materials and Methods.

### Antigen mapping and HLA assignment

The MIRA assay was used for antigen mapping as described previously (32). To assign HLA subtypes, ESs were clustered by a simple TCR amino acid similarity metric. ESs were considered to belong to a cluster if they shared a V-gene family and if all members of a cluster were connected by no more than a 1-Hamming difference between CDR3 regions. HLA subtypes were inferred based on the results of one-tailed FETs performed between each ES and every HLA subtype. For a detailed description of MIRA experiments and assignment of ESs to antigens and HLA subtypes, see the Supplementary Materials and Methods.

### Statistical analysis

Detailed statistical analyses associated with development of the TCR classifier and MIRA-based TCR–antigen assignment are described above. Additional statistical analyses were performed using the Python packages, scipy (version 1.5.4) and statsmodels (version 0.12.2). Significant associations between TCR score and clinical variables were assessed by the Mann-Whitney *U* test in univariate analysis and by multivariable linear regression with sex, age, and serostatus as variables. *P* values <0.05 were considered significant. The correlation between TCR score and the number of LD-related symptoms was assessed by Spearman’s rank-order correlation. For comparisons of sensitivity, error bars represent mean ± 95% CI by bootstrap sampling. For box-and-whisker plots, boxes indicate median ± interquartile ranges (IQR), and whiskers denote 1.5 times the IQR above the high quartile and below the low quartile.

### Data availability

Deidentified data are available by request from AdaptiveBiotechnologies Medical Information (https://www.adaptivebiotech.com/medical-information-request/).

## Data Availability

Data requests may be submitted for consideration to Adaptive Biotechnologies Medical Information (https://www.adaptivebiotech.com/medical-information-request/).

## Acknowledgments

We are grateful to the research participants who contributed samples and data used in this study and to the physicians/health care providers who facilitated recruitment.

Funding for this study was provided by Adaptive Biotechnologies. Institutional support was provided by the Steven and Alexandra Cohen Foundation (A.W.R., M.J.S., E.J.H., J.N.A.), Global Lyme Alliance (A.W.R., J.N.A.), National Institutes of Health grant P30 AR070254 (M.J.S.), and Bay Area Lyme Foundation (E.J.H.).

Medical writing and editorial support were provided by Melanie Styers and Rachel Salmon of BluPrint Oncology Concepts and Kristin MacIntosh and Shahin Shafiani of Adaptive Biotechnologies.

## SUPPLEMENTARY MATERIALS

### Materials and Methods

Figure S1. Immunosequencing input DNA distributions by cohort.

Figure S2. Clinical correlates of TCR scoring in STTT+ and STTT- individuals.

Figure S3. Receiver operating characteristic curves using all control samples from Lyme-endemic regions from Figure 2A as negatives.

Table S1. Multiple logistic regression of TCR model score on clinical features.

Table S2. Counts of enhanced sequences mapped to each protein by MIRA.

## SUPPLEMENTARY MATERIALS

### MATERIALS AND METHODS

#### TCR repertoire immunosequencing

Immunosequencing of TCRβ CDR3 was performed using the immunoSEQ^®^ Assay (Adaptive Biotechnologies, Seattle, WA, USA). Extracted genomic DNA was amplified in a bias-controlled multiplex PCR, followed by high-throughput sequencing. Sequences were collapsed and filtered to identify and quantitate the absolute abundance of each unique TCRβ CDR3 region for further analysis as previously described (26, 52, 53). Sequencing reactions contained a median of 7,884.0 ng of input DNA (range, 239.9–55,186.4 ng) and yielded a median of 314,948 T-cell templates per sample (range, 15–1,837,496) (Figure S1). The T-cell fraction (percentage of T cells among the estimated number of nucleated cells input) ranged from 0.3% to 90% (median, 26.0%).

#### TCR repertoire QC criteria

The 2 key parameters of the classifier are the number of unique productive rearrangements, *x*, and the number of unique productive rearrangements encoding an ES, *y*. For a given blood sample, the value of *x* is determined by the quantity of DNA, the fraction of cells that are T cells, and the diversity of T cells. In rare cases, *x* is too small to yield meaningful information, or significantly larger than observed in our training data, making extrapolation of *Y*|*x* problematic. Therefore, acceptance criteria were predefined for the number of unique productive rearrangements based on the observed distribution of *x* in the training data.

The information contained in ESs is asymmetric: for small *x*, large *y* is considered to be evidence of LD, while small *y* may simply reflect a lack of sequenced T cells. Thus, QC criteria were treated asymmetrically. Specifically, *x*_max_ and *x*_min_ were defined as the upper and lower QC thresholds, which were prespecified to be equal to the 1st and 99th percentiles, respectively, of *x* observed in the training data. A sample *i* then failed QC if *x*_*i*_ > *x*_max_ , or if *x*_*i*_ < *x*_min_ and *ModelScore*(*y*_*i*_, *x*_*i*_) < *t*.

#### Determination of antigen specificity of Lyme-associated TCRs

To evaluate the potential specificity of antigens recognized by TCRs in the LD classifier, ESs were clustered by sequence similarity. Statistical assignment of individual TCRs to HLA subtypes resulted in a consistent HLA assignment for the cluster. To further characterize TCR– antigen specificity, MIRA was used to identify TCR epitopes, as detailed in the next section.

Query peptides (777 total) were derived from 26 *Bb* proteins, and individual peptides or groups of related peptides were assigned to 1 of 426 unique MIRA pools. MIRA was then performed on T cells derived from peripheral blood mononuclear cells collected from 395 healthy individuals using a version of the assay that selects for HLA-II‒restricted CD4+ T cells. Basic Local Alignment Search Tool (BLAST) analysis of the target epitope was used to compare *Bb* FlaB to other *Borrelia* species and other pathogens. Specificity of our approach was further evaluated by comparing the Lyme-associated ESs with a set of TCRs from 507 individuals that were previously mapped to 325 SARS-CoV-2 antigen pools by MIRA (29 and Supplemental reference 1).

#### Antigen-stimulation experiments (MIRA assay)

##### Panel design

The multiplex identification of antigen-specific TCRs (MIRA) assay was set up, performed, and analyzed as described previously (32). Briefly, 2 panels of peptides were designed and tested in the assay. The Lyme-MIRA1 panel used antigens that are known to be presented in LD, including tiled portions of the DbpA, OspC, OspA, BBK32, BBA52, and VlsE proteins (Supplemental reference 2). The Lyme-MIRA2 panel used peptides derived from antigens presented via HLA class II upon *Bb* infection. The *Bb*-derived antigens include elongation factor Tu (WP_002657015.1), BB_0418 (WP_002658797.1), p83/100 (CAA57125.1), ABC transporter (PRR58667.1), lipoprotein LA7 (WP_002657819.1), GAPDH (AAB53930.1), chaperonin GroEL (WP_002657108.1), flagellin (WP_002661938.1), OspA (WP_010890378.1), and p66 (WP_002656762.1). The peptide tiling strategy was used across the entirety of each antigen, yielding a series of peptides, each 17 amino acids (aa) long with a 7-aa overlap between peptides.

The peptides were pooled in a combinatorial fashion as described previously (32); peptides that were overlapping or in close proximity in the viral proteome were grouped together into antigen sets. Each antigen set was then placed in a subset of 5 unique pools out of 11 total pools in the Lyme-MIRA1 panel, or 6 pools out of 12 in the Lyme-MIRA2 panel, referred to as its occupancy.

##### Naïve antigen-stimulation experiments

A total of 304 experiments were run with the Lyme-MIRA1 panel (all “naïve” experiments, see below) and 174 with the Lyme-MIRA2 panel. For the “naïve” experiments, CD14+ monocytes were selected from peripheral blood mononuclear cells (PBMCs; Miltenyi Biotech, Auburn, CA, USA) and stimulated with granulocyte-macrophage colony-stimulating factor (GM-CSF) and interleukin (IL)-4 (BioLegend, San Diego, CA, USA) to drive dendritic cell (DC) differentiation *in vitro*. On day 3, GM-CSF, IL-4, interferon-γ (IFN-γ; BioLegend, San Diego, CA, USA), and lipopolysaccharide (LPS; Sigma-Aldrich, St. Louis, MO, USA or eBioscience, Inc, San Diego, CA, USA) were added to promote DC maturation. Also on day 3, naïve T cells were isolated from PBMCs (StemCell, Vancouver, BC, Canada) and incubated overnight with IL-7. On day 4, naïve T cells were combined with the differentiated CD14+ monocytes, IL-21 (BioLegend, San Diego, CA, USA), and a pool of all peptides present in the panel to be used for restimulation.

Cultures were supplemented with IL-7, IL-15, and IL-2 every 2 to 3 days for an additional 12 to 14 days. Cells harvested from the expansion culture were divided into a series of replicate cultures, and each was restimulated using a distinct peptide pool from the panel under investigation. After incubation at 37°C for ∼20 hours, each culture was stained with antibodies (BioLegend, San Diego, CA, USA) for sorting by flow cytometry. Cells were then washed and suspended in phosphate-buffered saline containing 2% fetal bovine serum (FBS), 1 mM EDTA, and 4,6-diamidino-2-phenylindole (DAPI) for exclusion of non-viable cells. Cells were acquired and sorted using a FACSMelody (BD Biosciences, Franklin Lakes, NJ, USA) instrument.

Sorted antigen-specific (CD4+CD137+CD145+, CD25lo) T cells were pelleted and lysed in RLT Plus buffer (Qiagen, Germantown, MD, USA) for nucleic acid isolation.

##### Assignment of ESs to antigens

To assign ESs to antigens, RNA was isolated using AllPrep DNA/RNA mini and/or micro kits, according to the manufacturer’s instructions (Qiagen). RNA was then reverse transcribed to cDNA using Vilo kits (Life Technologies, Carlsbad, CA, USA), and TCRβ amplification was performed using the immunoSEQ Assay described above.

After immunosequencing, the behavior of T-cell clonotypes was examined by tracking read counts across each sorted pool. True antigen-specific clones should be specifically enriched in a unique occupancy pattern corresponding to the presence of 1 of the query antigens in 5 or 6 pools in the Lyme-MIRA1 and Lyme-MIRA2 panels. Methods used to assign antigen specificity to TCR clonotypes have been reported previously (32). In addition to these methods, a non- parametric Bayesian model was developed to compute the posterior probability that a given clonotype was antigen-specific. This model uses the available read counts of TCRs to estimate a mean-variance relationship within a given experiment, as well as the probability that a clone will have zero read counts due to incomplete sampling of low-frequency clones. Together, this model considers the observed read counts of a clonotype across all pools and estimates the posterior probability of a clone responding to all valid addresses and an additional hypothesis that a clone is activated in all pools (truly activated, but not specific to any of our query antigens). To define antigen-specific clones, we identified TCR clonotypes assigned to a query antigen from this model with a posterior probability ≥0.7.

TCR sequences from MIRA were compared to the ES list based on V-gene, J-gene, and CDR3 amino acid sequences. Any exact matches between the 2 lists, in which the MIRA TCR sequence was found in at least 2 separate individuals, were considered sufficient to map the ES to the MIRA antigen.

#### ES clustering and HLA inference

Clustering of ESs was based on TCR amino acid similarity. Specifically, 2 TCRs were assigned to the same cluster if they shared a V-gene family (and so have similar CDR1 and CDR2), had identical length, and differed by, at most, 1 amino acid in the CDR3 region. Clusters with at least 5 ESs were reported (Table 4). A sequence motif representing the CDR3 amino acid sequences assigned to each cluster was generated using WebLogo (University of California, Berkeley, CA, USA) (Supplemental references 3, 4).

To assign an ES to a single HLA subtype, a one-tailed FET was performed between that ES and every HLA subtype. The ES was assigned to the HLA subtype with the lowest *P* value; if the lowest *P* value was >0.001, no assignment was made. Contingency tables counted the number of individuals with/without the ES and with/without a given HLA subtype. For HLA-DQ and HLA-DP, α/β heterodimers were treated as distinct HLA subtypes; for example, individuals with 2 α subtypes and 2 β subtypes were treated as expressing all 4 possible heterodimers. An HLA subtype was assigned to an ES cluster if most (>50%) of the cluster members with an assigned HLA subtype were assigned to the same subtype.

**Figure S1.**
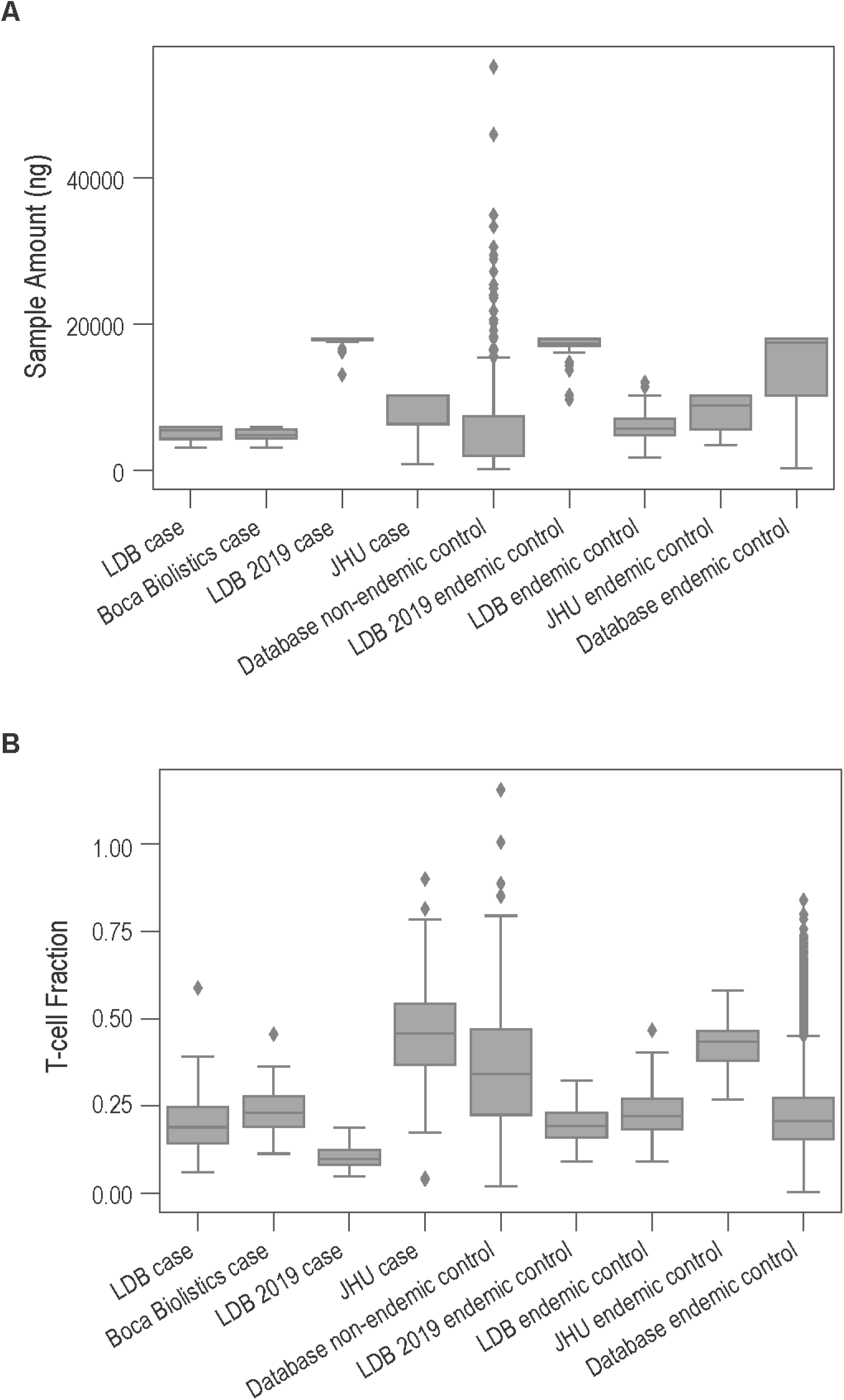
Immunosequencing input DNA distributions by cohort. Boxes indicate median ± IQR, and whiskers denote 1.5 times the IQR above the high quartile and below the low quartile.

**Figure S2.**
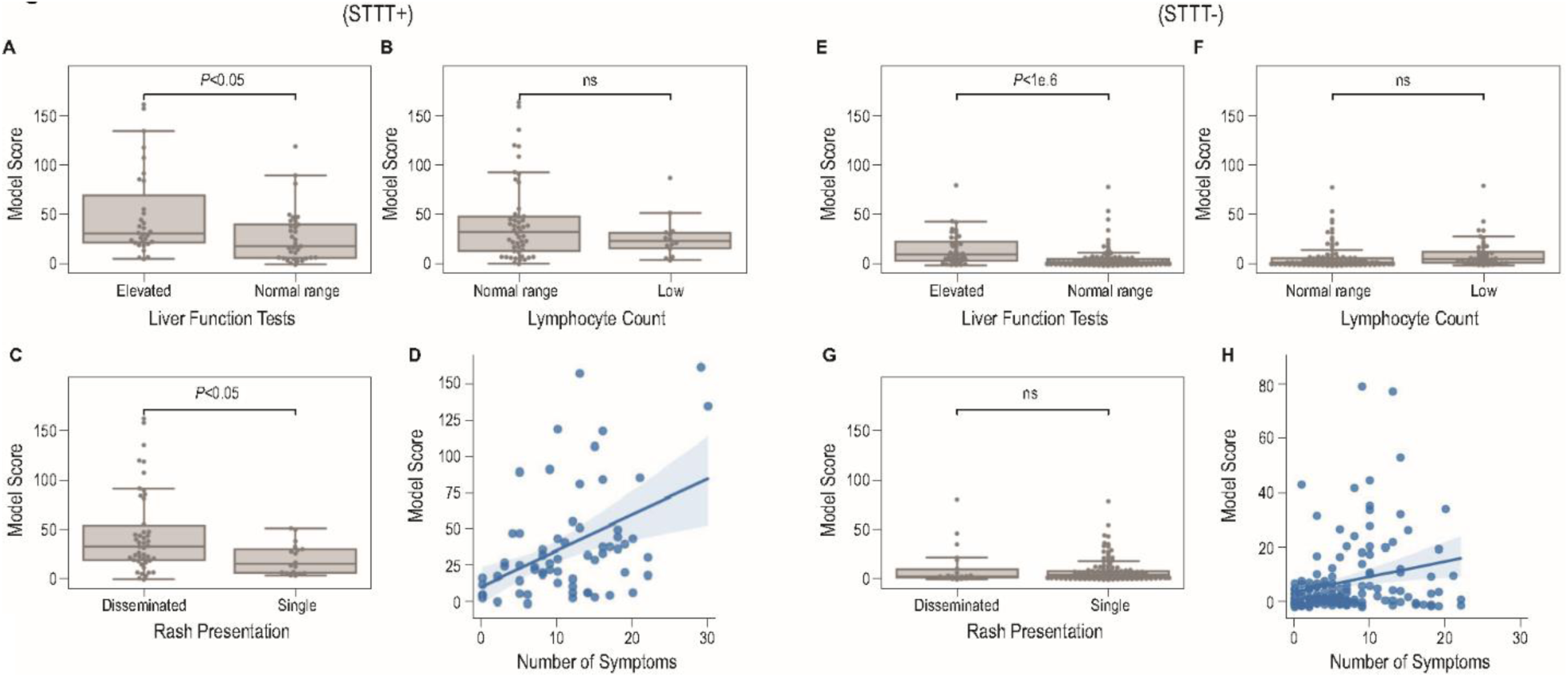
Clinical correlates of TCR scoring in STTT-positive (A–D) and STTT-negative (E–H) individuals. TCR scores were stratified by (**A, E**) liver function test results (elevated [n=72] vs normal [n=139]), (**B, F**) lymphocyte counts (normal [n=150] vs low [n=61]), or (**C, G**) presentation of rash (multiple/disseminated [n=68] vs single [n=143]) and (**D, H**) plotted as a function of the number of Lyme-related symptoms (Spearman R^2^=0.17. *P* values, Mann-Whitney *U* test). Ns, not significant. In box-and-whisker plots, boxes indicate median ± IQR, and whiskers denote 1.5 times the IQR above the high quartile and below the low quartile.

**Figure S3.**
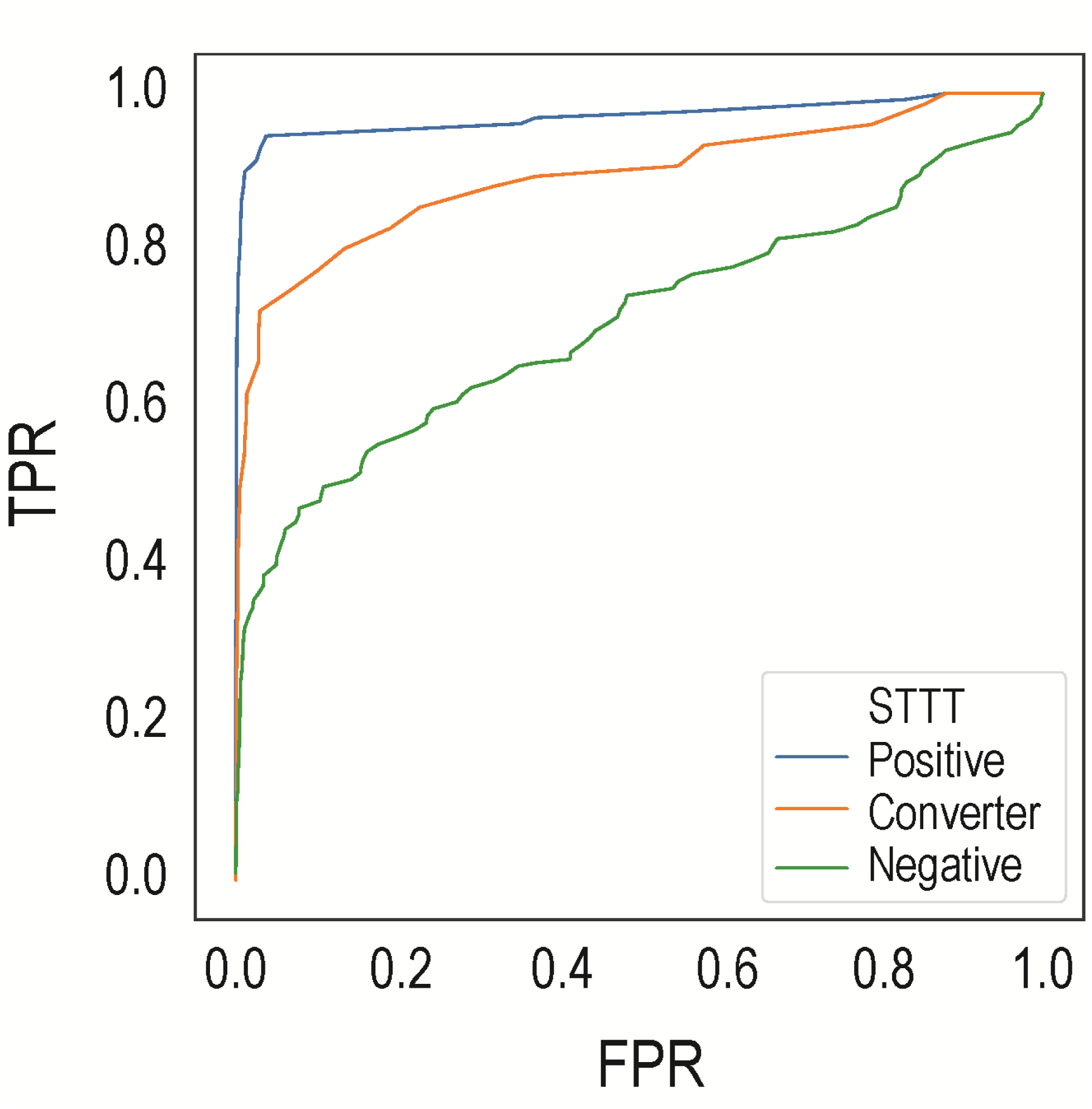
Receiver operating characteristic curves using all control samples from Lyme-endemic regions from Figure 2A as negatives. Areas under the receiver operating characteristic curves are 0.98, 0.89, and 0.71 for the positive, converter, and negative curves, respectively. FPR, false positive-rate; TPR, true-positive rate.

**Table S1.** Multiple logistic regression of TCR model score on clinical features.

|  | **Coef** | **SE** | **t** | ***P*>\|t\|** | **[0.025** | **0.975]** |
| --- | --- | --- | --- | --- | --- | --- |
| Intercept | -4.4736 | 7.323 | -0.611 | 0.542 | -18.914 | 9.966 |
| STTT (positive) | 17.1930 | 4.177 | 4.116 | <0.001 | 8.956 | 25.430 |
| Liver function tests (elevated) | 11.7845 | 3.537 | 3.332 | 0.001 | 4.809 | 18.760 |
| Local/disseminated rash (disseminated) | 11.6126 | 4.020 | 2.888 | 0.004 | 3.685 | 19.541 |
| Lymphocyte count category (normal range) | 5.5743 | 3.559 | 1.566 | 0.119 | -1.444 | 12.593 |
| Sex (male) | 2.0499 | 3.260 | 0.629 | 0.530 | -4.378 | 8.478 |
| Number of symptoms | 1.0257 | 0.282 | 3.633 | <0.001 | 0.469 | 1.582 |
| Rash area (mm<sup>2</sup>) | 0.0080 | 0.012 | 0.644 | 0.521 | -0.017 | 0.033 |
| Days from symptom onset to sample | -0.1968 | 0.184 | -1.067 | 0.287 | -0.561 | 0.167 |
| Age | -0.0866 | 0.102 | -0.847 | 0.398 | -0.288 | 0.115 |

**Table S2.** Counts of enhanced sequences mapped to each protein by MIRA.

| **Protein (Antigen)** | **No. of Enhanced Sequences** | **Total No. of Matches Across Experiments** |
| --- | --- | --- |
| FlaB (A) | 7 | 99 |
| FlaB (B) | 1 | 5 |
| DbpA | 1 | 4 |
| **Total** | **9** | **108** |

